# Comparison of media and standards for SARS-CoV-2 RT-qPCR without prior RNA preparation

**DOI:** 10.1101/2020.08.01.20166173

**Authors:** Katherine B. Ragan, Sanchita Bhadra, Joon H. Choi, Dalton Towers, Christopher S. Sullivan, Andrew D. Ellington

## Abstract

Since the emergence of the severe acute respiratory syndrome coronavirus 2 (SARS-CoV-2) pandemic, there have been demands on the testing infrastructure that have strained testing capacity. As a simplification of method, we confirm the efficacy of RNA extraction-free RT-qPCR and saline as an alternative patient sample storage buffer. In addition, amongst potential reagent shortages, it has sometimes been difficult to obtain inactivated viral particles. We have therefore also characterized armored SARS-CoV-2 RNA from Asuragen as an alternative diagnostic standard to ATCC genomic SARS-CoV-2 RNA and heat inactivated virions and provide guidelines for its use in RT-qPCR.

## Introduction

The causative agent of COVID-19 is the highly contagious severe acute respiratory syndrome coronavirus 2 (SARS-CoV-2). Due to the long asymptomatic incubation period of SARS-CoV-2 infections and the high percentage of infected individuals that are non-symptomatic, mass testing is necessary to contain the spread of the virus. One of the standard tests for SARS-CoV-2 is extraction and purification of RNA from patient nasal swabs preserved in viral transport media (VTM) or saline followed by quantitative reverse transcription PCR (RT-qPCR) to identify the presence of viral RNA in samples. However, RNA extraction is both time consuming and costly, and in order to increase testing capacity it may prove useful to attempt RNA-extraction free RT-qPCR. To this end, we attempted to look at the impact of different media and different positive control standards that might prove useful in modified assays. Overall, our data further demonstrate that no-RNA prep, direct RT-qPCR can be reproducibly used for viral RNA detection, albeit with lower sensitivity, and that saline is a superior alternative to VTM for collection. Importantly, we provide useful guidelines for using commercial armored RNA standards rather than inactivated virus as a positive control in assays.

## Results and Discussion

### Omitting RNA purification can allow high-throughput RT-qPCR diagnosis of SARS-CoV-2

As there is an increased need for testing for SARS-CoV-2 in the population, higher throughput methods for analysis are gaining sway. In particular, while RT-qPCR remains the gold standard for analysis, the requirement of RNA purification prior to enzymatic amplification slows the overall throughput of the assay and limits the number of samples that can be tested. In consequence, many groups have reported on “direct RT-PCR” assays for CoV-2 RNA that bypass the RNA purification^1–19^. As a prelude to understanding sample preparation and controls for assay development, we similarly assessed the sensitivity of an extraction-free RT-qPCR assay for naked genomic SARS-CoV-2 RNA (ATCC). Naked RNA was serially diluted in saline and directly subjected to RT-qPCR without RNA extraction, and could be detected at a concentration of 50 copies per 20 microliter qPCR reaction (approximately 2.5 copies per microliter) with the N1 primer set, which amplifies the nucleocapsid (N) gene of SARS-CoV-2 (**Figure 1**), confirming extraction-free RT-qPCR as a plausible diagnostic tool^1-19^.

**Figure 1:**
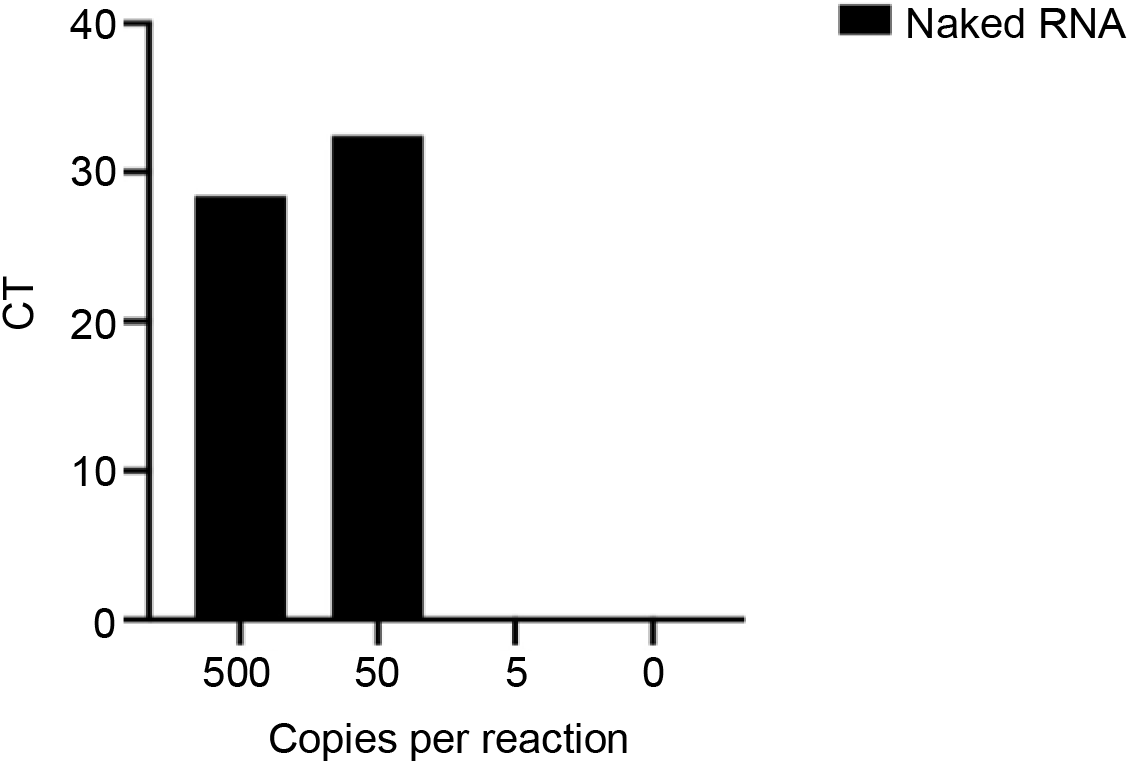
Omitting RNA purification can allow high-throughput RT-qPCR diagnosis of SARS-CoV-2. RNA extraction-free RT-qPCR assessing the sensitivity of detection of naked SARSCoV-2 RNA diluted in saline using the N1 primer/probe set.

### Benchmarking armored SARS-CoV-2 RNA as a diagnostic standard

Since genomic SARS-CoV-2 RNA standards have become limited in availability (ATCC has experienced backorders; BEI provides limited samples), we also used this opportunity to validate a second standard, armored SARS-CoV-2 RNA (Asuragen, Austin, TX). Armored RNAs are generated by including a phage packaging sequence on a given RNA standard, which leads to its being coated in phage MS2 coat protein. This in essence creates a pseudo-virion that is likely less susceptible to RNase degradation than uncoated, naked RNA.

We diluted armored RNA directly into saline and found it could be detected without additional preparation at comparable levels to naked RNA with the N1 primer set at concentrations of 500 copies per reaction and 50 copies per reaction; **Figure 2A**). However, the sensitivity of the assay with protein-coated RNA was less than that of naked RNA (CT value of 35.9 at 50 copies/reaction, compared with CT value of naked RNA of 32.5 at the same concentration; **Figure 2A**). We further assessed the efficacy in direct RT-qPCR of the N2 and N3 primer/probe sets, which amplify different regions of SARS-CoV-2’s nucleocapsid gene. It should be noted that the N1 and N2 primer/probe sets have passed functional testing by the CDC while the N3 set has not. While both the N1 and N3 primer/probe sets efficiently detected both naked and armored RNA in our direct assay, yielding an average CT value of 32.6 for naked RNA and 35.3 for armored RNA at a concentration of 50 genomic copies per reaction, the N2 primer/probe set only amplified naked RNA at a concentration of 500 genomic copies per reaction (**Figure 2B**).

**Figure 2:**
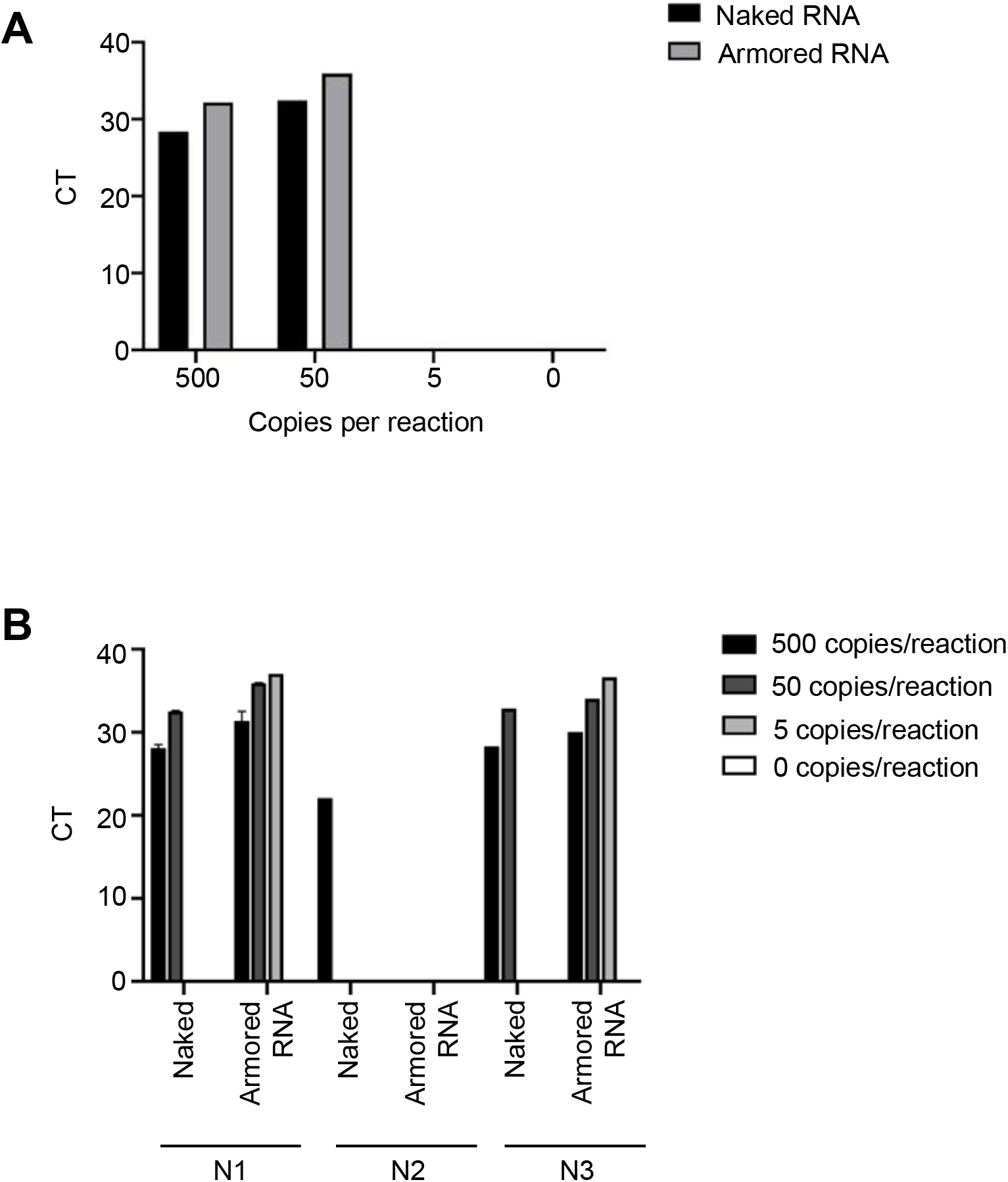
Armored SARS-CoV-2 RNA is a viable alternative diagnostic standard. A) RNA extraction-free RT-qPCR assessing the sensitivity of detection of naked SARSCoV-2 RNA compared to armored SARS-CoV-2 RNA diluted in saline using the N1 primer/probe set. B) RT-qPCR assessing the efficacy of the N1, N2, and N3 primer/probe sets in an RNA extraction free setting in amplifying naked SARS-CoV-2 RNA and armored SARS-CoV2 RNA diluted in saline. Error bars represent standard deviation for n = 2 for the N1 primer/probe set data.

### Dilution in viral transport media reduces the sensitivity of detection

Current CDC standards recommend that nasal swabs be stored in viral transport media (VTM) or, if VTM is unavailable, sterile saline. VTM is a solution that was developed for culturing viral samples rather than for preserving viral RNA for detection by RT-qPCR. We hypothesized that since VTM is a complex media that contains heat inactivated fetal bovine serum, gentamicin, and fungizone (amongst other additives), one or more components might act as an interferent in RT-qPCR. To best simulate real world clinical samples stored in VTM, four negative patient samples collected in VTM were combined at equal ratios to generate what will henceforth be referred to as pooled patient-negative VTM. An additional standard, heat inactivated SARS-CoV-2 virions, was added to the assay to mimic infected patient samples. Naked and armored RNA were also diluted into the pooled patient-negative VTM.

While naked RNA and heat inactivated virions were both detectable in extraction-free RT-qPCR in saline at 5 copies per reaction under our assay conditions, naked RNA was undetectable in pooled patient-negative VTM. This was perhaps unsurprising, given the likely prevalence of RNases in patient samples. The detection of armored RNA was less sensitive than the detection of inactivated CoV-2 viral particles (50 copies per reaction for armored RNA in VTM, versus 5 copies for inactivated virions; **Figure 3A**). Assays with both armored RNA and heat inactivated viral particles were slightly less sensitive when diluted in VTM than in saline, with slight increase in CT values of (averaging 2.9) using the N1 primer/probe set (**Figure 3A**). Together, these data are consistent with direct dilution in saline being preferable to direct dilution from VTM and confirm others’ observations that saline can serve as an effective sample storage buffer^3,8,20,21^

Because pooled patient-negative VTM was less conducive to the amplification of naked RNA under our assay conditions, we wanted to see if this decrease in sensitivity could be ameliorated by diluting pooled patient-negative VTM into saline. To test this, 1 microliter of pooled patient-negative VTM was added to naked and armored RNA diluted in saline, and amplification using the N1 primer/probe set was compared to samples diluted in either water or pure saline. Naked RNA does amplify in the presence of diluted pooled VTM at concentrations of 500 and 50 copies/reaction, but with an average increase in CT value of 7.2, compared with naked RNA diluted in pure saline. (**Figure 3B**).

### The impact of RNases on amplification

To better parse out the role of residual RNases in assay inhibition, we assessed the amplification of each standard using the N1 primer/probe set on a negative patient swab stored in saline (referred to as patient-negative saline) and compared it to amplification of RNA in pure saline and pooled patient-negative VTM. Surprisingly, the patient-negative saline supported amplification of naked RNA, with a small detrimental effect (an increase in CT value of 2.4 compared with pure saline; **Figure 3A**). In contrast, the change in CT values for both armored RNA and heat inactivated virions is negligible when diluted in patient negative saline versus pure saline: armored RNA has an increase in CT value of 0.4, while the CT value for heat inactivated virions actually slightly decreases when amplified in patient negative saline (**Figure 3A**). Overall, these results would seem to suggest that RNases play only a small role in inhibiting amplification following direct dilution, and that components of VTM have a more substantive effect. That said, we note that our patient-negative saline sample was harvested from one individual, and that assays were carried out immediately following the addition of templates in any form, which may have limited the activity of endogenous RNases.

To further examine the potential impact of RNases on amplification, we added SUPERase-in RNase inhibitor to samples stored in pooled patient-negative VTM. Armored RNA was diluted in saline, water, pooled patient-negative VTM diluted in saline, and patient-negative saline and assayed using the N3 primer/probe set with or without SUPERase-in. Under the conditions we utilized (spiking 1 uL into the RT-qPCR reaction) SUPERase-in addition was actually found to increase the CT values of armored RNA (**Figure 3C**).

**Figure 3:**
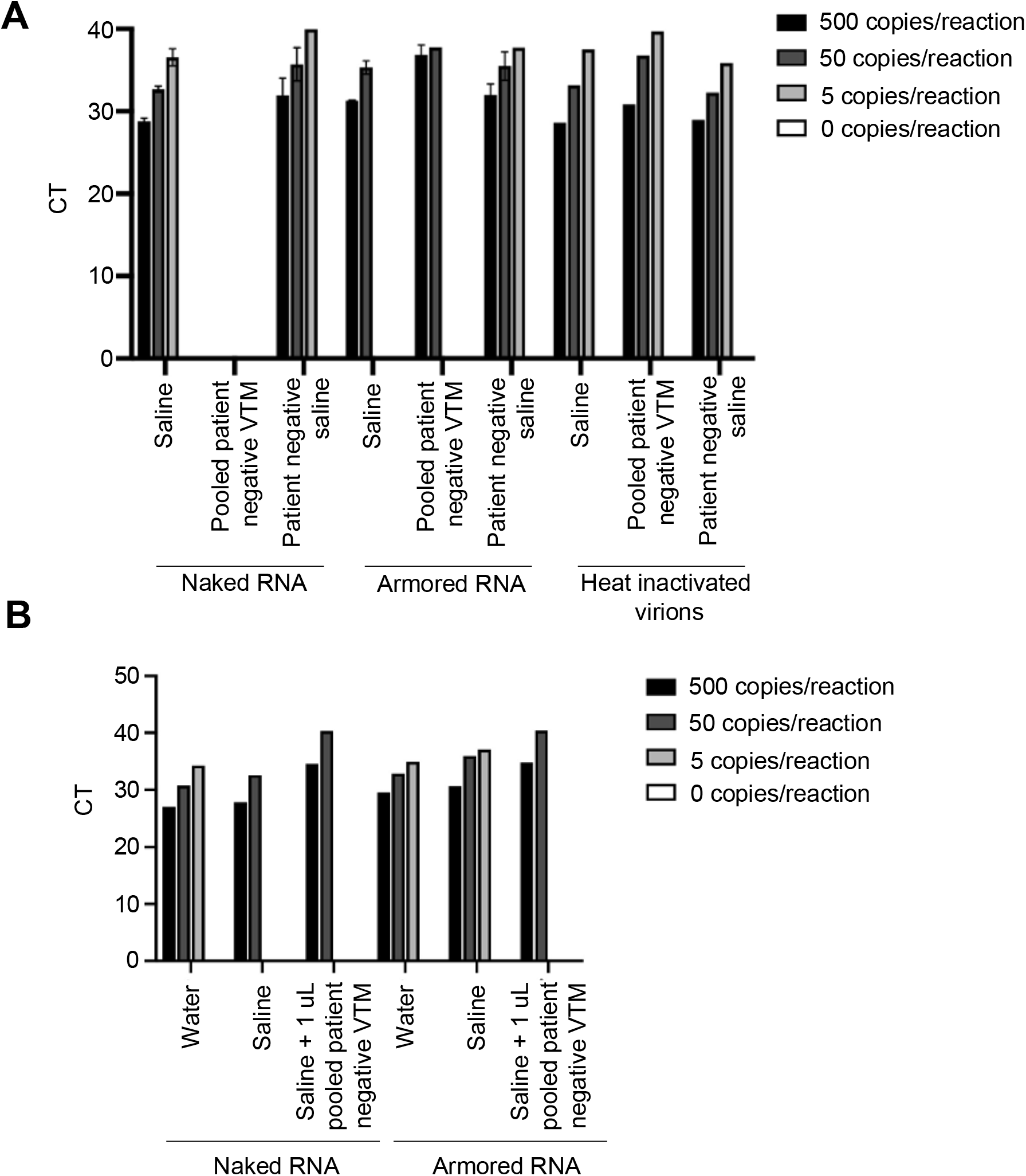
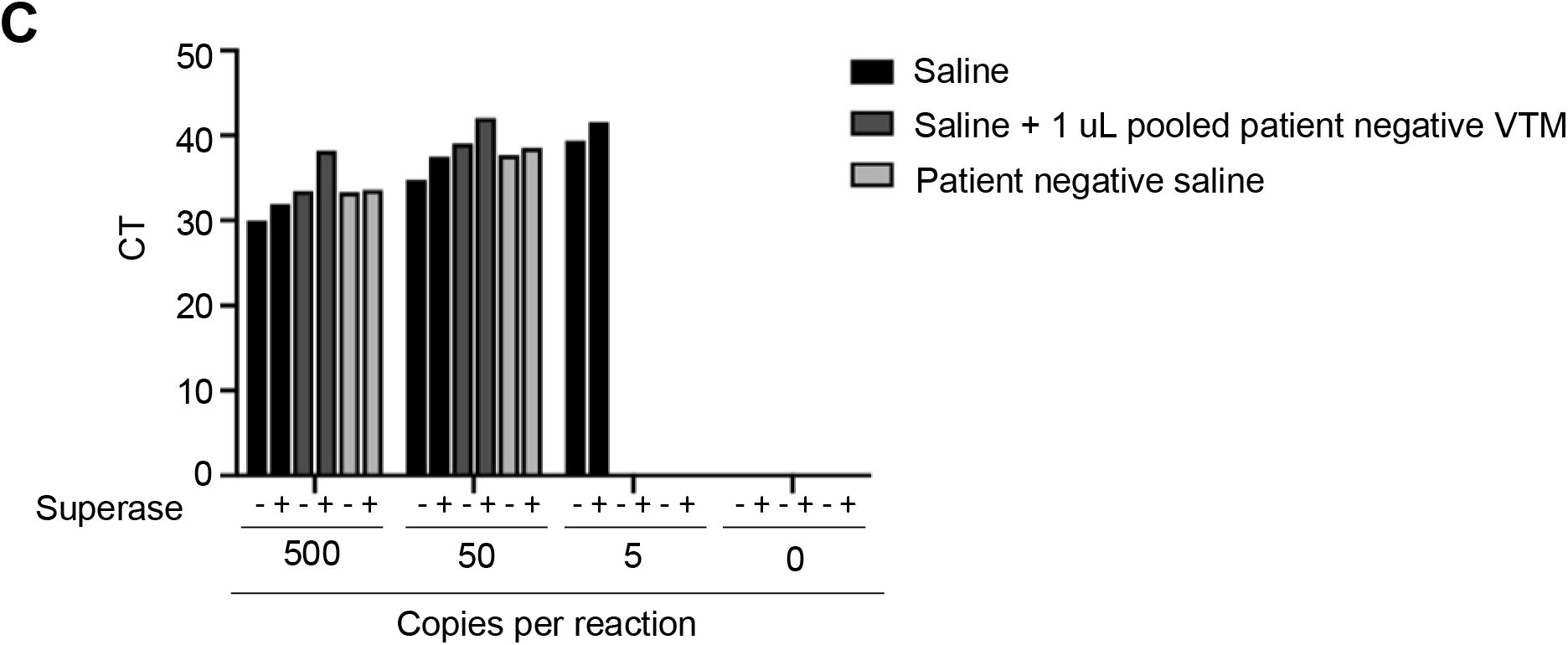
Extraction-free RT-qPCR is more sensitive at detecting SARS-CoV-2 RNA when samples are diluted in saline versus viral transport media. A) RNA extraction-free RT-qPCR comparing amplification of naked SARS-CoV-2 RNA, armored SARS-CoV-2 RNA, and heat inactivated SARS-CoV-2 virions in saline, pooled patient-negative VTM, or patient-negative saline using the N1 primer/probe set. B) RNA extraction-free RT-qPCR comparing amplification of naked SARS-CoV-2 RNA to armored SARS-CoV-2 RNA in water, saline, or saline plus 1 uL of pooled patient-negative VTM using the N1 primer/probe set. C) RNA extraction-free RT-qPCR comparing amplification armored SARS-CoV-2 RNA, in saline, saline plus 1 uL of pooled patient-negative VTM, or patient negative saline in the presence or absence of SUPERase-in RNase inhibitor using the N3 primer/probe set.

### Treating armored RNA for enhanced detection

Having noted that detection of armored RNA seems less sensitive than the detection of naked viral RNA or inactivated capsids (**Figure 3A**), we attempted to determine if there were conditions that would improve armored RNA detection. The manufacturer suggests a heat treatment of 75 C for 3-5 min should be sufficient to uncoat the RNA. We therefore compared the effect of two heat treatments on the amplification of armored RNA. The recommended heat treatment at 75 C for 5 minutes does not seem to improve amplification of armored RNA compared to untreated samples, with an average increase in CT value of 0.1 (**Figure 4A**). A more rigorous heat treatment of 65 C for 15 minutes followed by a 3 min incubation at 98 C actually had a slight negative effect on amplification compared with unheated samples, with an average increase in CT value of 0.9 across a dilution series (**Figure 4A**). Together these data suggest that heat treatment does not increase the sensitivity of extraction-free RTqPCR assay for armored RNA and in fact, can have a minor detrimental effect on detection.

We then attempted a brief treatment of armored RNA samples with Proteinase K, a serine protease that is often used for general protein digestion due to its broad substrate specificity^22^. A prepared dilution series of armored RNA was spiked with Proteinase K to a final concentration of 100 μg/mL followed by heat treatment at 65 °C for 15 minutes and a 3 min incubation at 98 °C to inactivate the enzyme prior to being loaded into RT-qPCR. Both untreated and Proteinase K treated samples were subjected to the heat treatment. As shown in **Figure 4B**, Proteinase K pretreatment actually increased the average CT value across the dilution series by 0.9. Reduction of sensitivity can potentially be explained by incomplete deactivation of Proteinase K.

**Figure 4:**
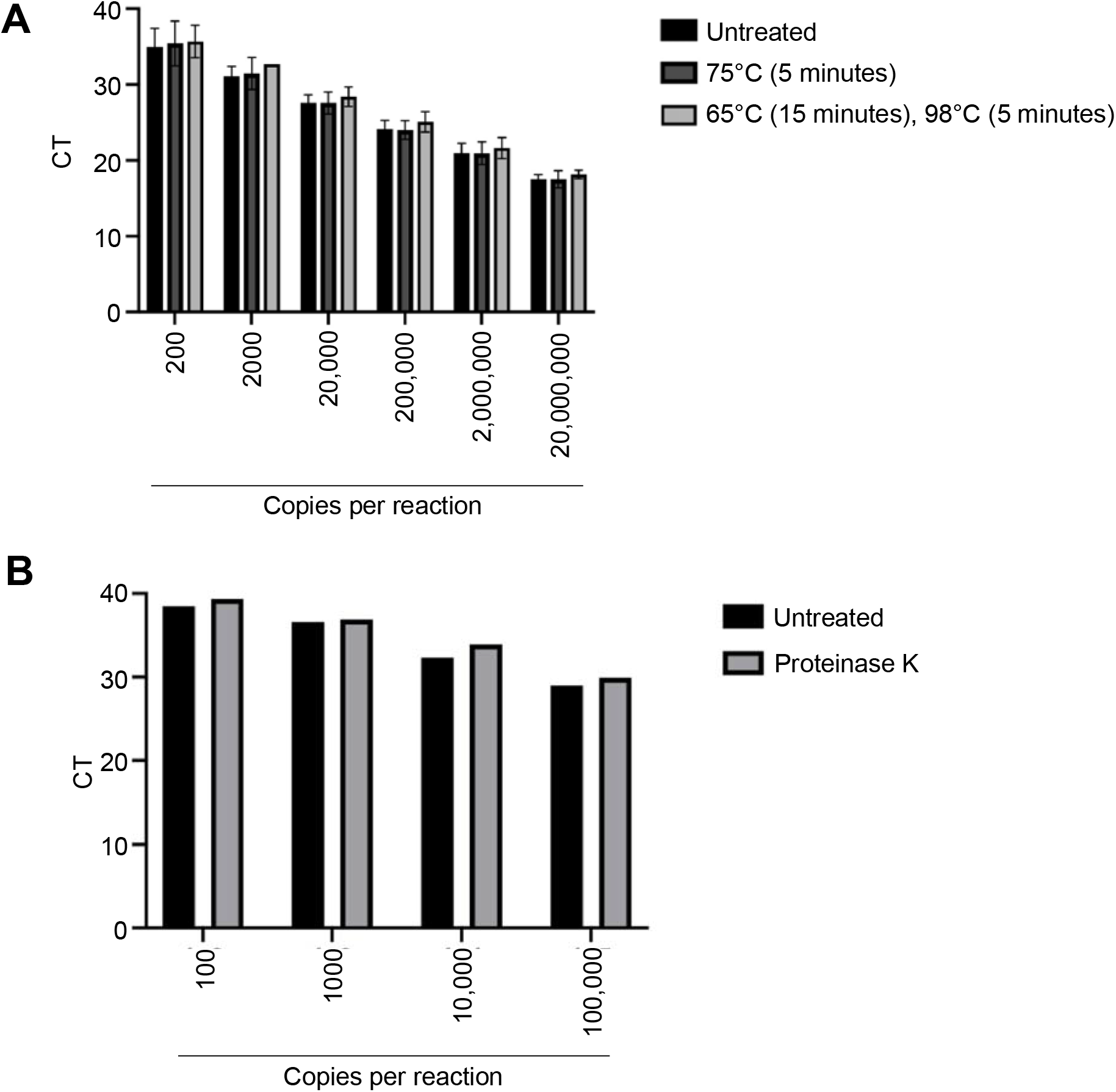
Treating armored RNA for enhanced detection. A) RNA extraction-free RT-qPCR comparing the effect of heat treatment on amplification of armored SARS-CoV-2 RNA. Samples diluted in 0.5 mM EDTA were then either untreated, heated at 75°C for 5 minutes, or heated at 65°C for 15 minutes followed by 98°C for 5 minutes. RT-qPCR was performed using the N1 primer/probe set. B) RNA extraction-free RT-qPCR assessing the effect of Proteinase K treatment (100ug/reaction) on amplification of armored RNA across a dilution series in 0.5 mM EDTA. RT-qPCR was performed using the N1 primer/probe set.

## Conclusions

Our work confirms previously published results showing that extraction-free RT-qPCR diagnosis of SARS-CoV-2 is possible^1-19^. Additionally, our results further support saline as a viable alternative to VTM for patient sample storage^3,8,20,21^. In comparison to VTM, saline affords greater sensitivity for detection of genomic SARS-CoV-2 RNA, possibly due to interferants present in VTM as well as reduced RNase activity in saline. Surprisingly, RNases that may have been present in patient samples appeared to have little effect on the sensitivity of amplification, at least when samples are immediately assayed via RT-qPCR following dilution. It remains possible that if samples were incubated at room temperature over a time course that the sensitivity of the RT-qPCR assay would decrease due to RNase activity. Far more significant is the impact of VTM on amplification, and it seems likely that one or more components of VTM inhibits either the reverse transcription or amplification steps. Since VTM is both costly and sometimes of limited availability, the use of saline for higher-throughput, no-RNA-prep, direct-dilution RT-qPCR assays is recommended.

We also find that armored RNA is a potentially useful standard for RT-qPCR. However, it seems to both protect and / or uncoat RNA templates in a way that is different than the natural virion. Heat-inactivated virions were consistently more detectable in extraction-free RT-qPCR than armored RNA, across all buffers and at all concentrations. Attempts to improve the limits of detection via heating armored RNA prior to amplification or a brief treatment with proteinase K did not lead to an increase in sensitivity. One possibility is that that the phage coat is generally more robust than the SARS-CoV-2 capsid. In addition, it should be noted that the methods used to quantify naked RNA in the patient-derived samples (spectrophotometric, versus the NIST-standardized phosphate assay performed by the manufacturer) and the way in which the standards were diluted (**0.5 mM EDTA**, versus the recommended TSMIII) may have impacted the quantitation differences observed. Thus, when using armored RNA to develop standards and new methods for RT-qPCR, it is important to keep in mind that determining sensitivity for this standard may require comparative quantitation, as we have carried out here.

## Materials and Methods

Genomic viral RNA (ATCC catalog #VR1986D), armored viral RNA (obtained from Asuragen), and heat inactivated viral particles (ATCC catalog #VR1986HK) were serially diluted to 10^3^ copies/mL in nuclease free water. From this concentration, subsequent working dilutions of 100, 10, and 1 copies/uL were made in the indicated buffers. The .9% saline solution was made by dissolving 9g NaCl into 1L of diH2O and autoclaved to sterilize. Viral transport media (VTM), patient-negative samples in VTM, and negative patient samples in saline were provided by Clinical Pathology Laboratories (CPL). Pooled patient-negative VTM was generated by combining 4 different patient-negative swabs stored in VTM at an equal ratio. Saline plus pooled patient-negative VTM was made by adding 1 uL of pooled patient-negative VTM to each qPCR reaction. Patient-negative saline was generated from one negative patient swab stored in saline. Where indicated, 1 uL of SUPERase-in RNase inhibitor (ThermoFisher AM2694) was spiked into each RT-qPCR reaction. Samples were not subjected to an RNA extraction step; rather, samples were directly assayed. Each sample was analyzed in duplicate using the TaqPath 1-step RT-qPCR GC master mix (Thermo catalog #A15300) on a VIIA 7 Real Time PCR machine using a MicroAmp^TM^ Fast Optical 96-well reaction plate. The N1, N2, and N3 primer/probe sets were obtained from IDT (catalog #1006606). The reaction set up is described in Table 1, and the cycling conditions are described in Table 3, below. Figure 1A, 1B, 2A, and 2B use some of the same data as the assays were done simultaneously.

**Table 1:** Reaction set up for Figures 1-3 experiments:

|  | Volume (uL) |
| --- | --- |
| Sample | 5 |
| Primer/probe mixture | 1.5 |
| TaqPath master mix (4x) | 5 |
| Nuclease free water | 8.5 |
| Total reaction volume | 20 |

For the experiments probing the effect of heat treatment and Proteinase K treatment on detection in extraction-free RT-qPCR, armored RNA was serially diluted as indicated into 0.5 mM EDTA into a final volume of 20 uL. Armored RNA was stored at -80 °C, as opposed to the -20 °C recommended by the manufacturer, and it is unclear how this impacted its eventual function. For Figure 4A, samples were either untreated, heated for 5 minutes at 75°C, or heated for 15 minutes at 65°C then 5 minutes at 98°C. For Figure 4B, samples were either untreated or treated with 100 ug of Proteinase K (Thermoscientific, EO0492) followed by heating for 15 minutes at 65°C then 5 minutes at 98°C. Samples were directly assayed in duplicate as described in Table 2, below, using the N1 primer/probe set and the TaqPath 1-step RT-qPCR GC master mix on a VIIA 7 Real Time PCR machine using a MicroAmp^TM^ Fast Optical 96-well reaction plate.

**Table 2:** Reaction set up for Figure 4 experiments:

|  | Volume (uL) |
| --- | --- |
| Sample | 20 |
| N1 Primer/probe mixture | 3 |
| TaqPath master mix (4x) | 10 |
| Nuclease free water | 7 |
| Total reaction volume | 40 |

**Table 3:** cycling conditions:

| Step | Duration | Temperature |
| --- | --- | --- |
| 1 | 2 minutes | 25°C |
| 2 | 15 minutes | 50°C |
| 3 | 2 minutes | 95°C |
| 45 cycles of steps 4 and 5 |  |  |
| 4 | 3 seconds | 95°C |
| 5 | 30 seconds | 55°C |

**Table 4:**
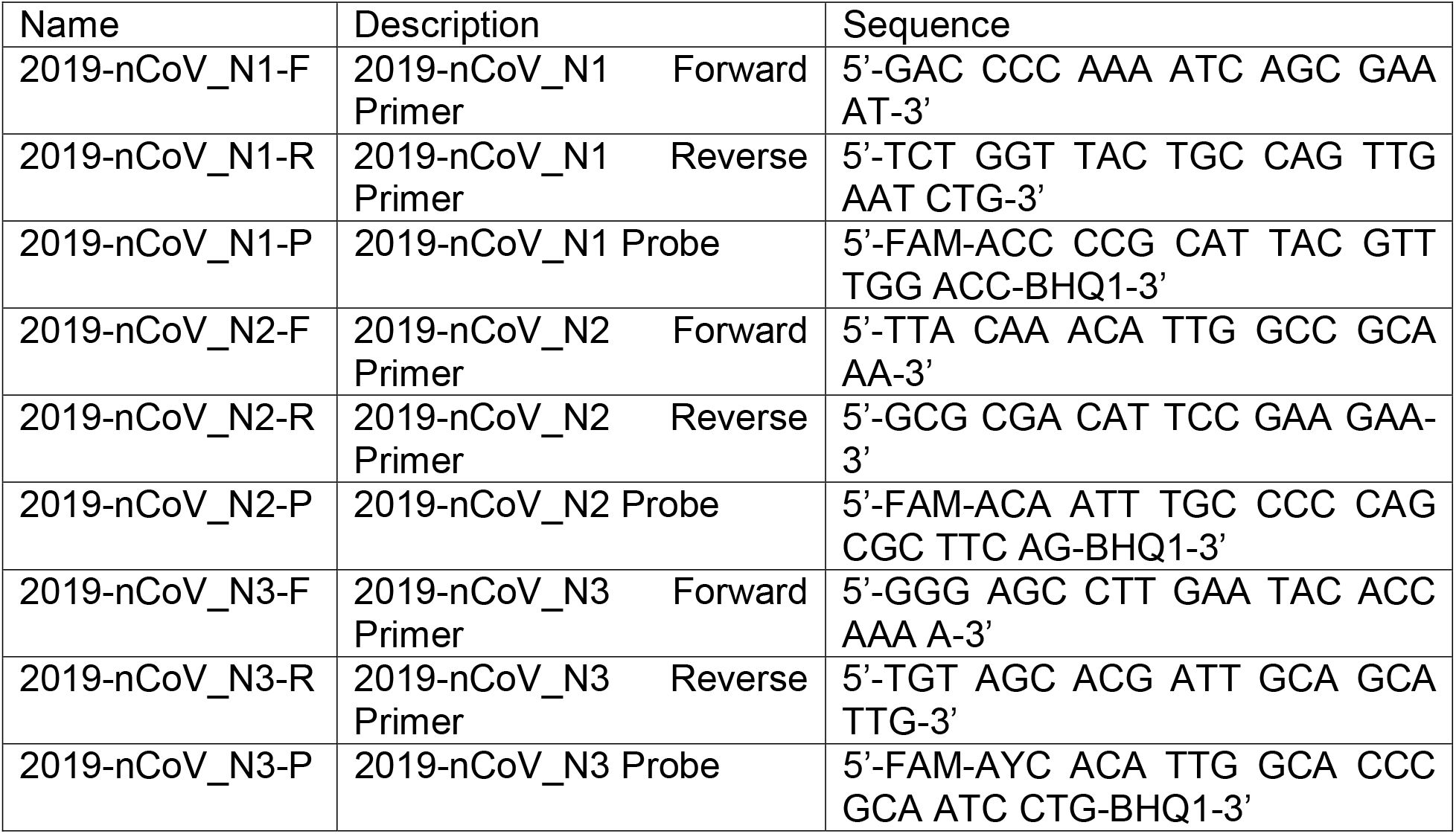
Primer and probe sequences

## Data Availability

There are no external data files.

## Acknowledgements

The authors thank Dr. Scott Hunicke-Smith at Clinical Pathology Laboratories for kindly providing us with patient-negative samples and Viral Transport Media for us to carry out this work as well as helpful input and insightful discussion. Work from the Ellington lab was supported by funding from the National Institutes of Health (1R01EB027202-01A1), the National Science Foundation (2027169), and the Welch Foundation (F-1654). Work from the Sullivan lab was supported by funding from the University of Texas at Austin Office of the Vice President for Research, the National Institutes of Health (R01AI123231 and 1R01AI134980), and a Burroughs Wellcome Investigators in Pathogenesis Award (1011070).

